# Exposure to Lead and Incidence of Alzheimer Disease and All-Cause Dementia in the United States

**DOI:** 10.1101/2025.08.15.25333790

**Authors:** Xin Wang, Kelly M. Bakulski, Erika Walker, Bhramar Mukherjee, Hiroko Dodge, Roger L. Albin, Henry L. Paulson, Sung Kyun Park

## Abstract

**INTRODUCTION:** Growing evidence suggests lead exposure may increase dementia risk, but evidence from human studies is limited. We investigated associations between lead exposure and incident Alzheimer disease (AD) and all-cause dementia in a nationally-representative, prospective cohort.

**METHODS:** We examined over 14,000 individuals with baseline measured blood lead and estimated patella and tibia lead from the National Health and Nutrition Examination Survey (NHANES)-III (1988-1994) and continuous NHANES (1999-2016), linked to Medicare and the National Death Index for incident AD and all-cause dementia, with up to 30 years of follow-up. Survey-weighted Cox regressions estimated hazard ratios (HRs) and 95% confidence intervals (CIs).

**RESULTS:** In continuous NHANES, estimated patella lead was positively associated with AD (HR=2.96, 95% CI: 1.37-6.39) and all-cause dementia (HR=2.15, 95% CI: 1.33-3.46), comparing quartile-4 vs. quartile-1. We observed weaker associations in NHANES-III. Blood lead showed no association.

**DISCUSSION:** These findings suggest cumulative lead as a potential dementia risk factor.

## 1. Introduction

Dementia, including Alzheimer disease (AD), represents a significant global health challenge, affecting millions worldwide and imposing substantial burdens on healthcare systems.^1^ The etiologies of AD and dementia are complex, involving a interplay of genetic and environmental factors.^2^ Despite considerable progress in understanding AD and dementia, significant knowledge gaps persist, especially regarding the influence of environmental factors on disease incidence. Among potential factors, the role of environmental toxicants, particularly lead, gained attention due to lead’s neurotoxic potential and widespread exposures.^3^

Lead toxicity is a prevalent public health issue, with common exposure pathways including air, dust, water, and food. Preclinical model evidence linked lead exposure to AD hallmark pathologies, including the promotion of formation and spread of amyloid-beta (Aβ) plaque and tau neurofibrillary tangles.^4^ Lead neurotoxicity extends to oxidative stress, neuronal apoptosis, and neuroinflammation.^5^ Though several studies examined lead’s association with cognitive decline,^6^ direct epidemiological evidence supporting lead exposure impacts on incident AD and all-cause dementia is limited.

A major methodological challenge in evaluating the chronic effects of long-term cumulative lead exposure in human epidemiologic studies is the lack of biologically relevant exposure biomarkers. Most epidemiologic studies rely on blood lead levels measured at a single time point as the primary indicator of exposure. Blood lead, which has a half-life of approximately 30 days, reflects mostly circulating lead from recent external exposure and the mobilization of lead from internal body stores.^7^ In contrast, bone lead, which serves as an indicator of cumulative lead exposure, has been recognized as a superior biomarker for assessing chronic effects due to its long elimination half-life, spanning years to decades.^7^ Reliance on blood lead alone introduces potential measurement error, potentially resulting in underestimation of the true effects of lead in risk assessments.^8^ Another key limitation of prior epidemiologic studies is the lack of incident dementia cases, as most have examined prevalent cases or mortality. Potential associations between lead exposure and the incidence of AD and all-cause dementia, particularly in a prospective cohort study setting, have not been thoroughly investigated.

This study aims to address these gaps by investigating measured blood lead as well as estimated bone lead variables linked to Medicare claims and the National Death Index for incident AD and all-cause dementia in a large, prospective cohort study. We hypothesize that higher exposure to lead is associated with increased risk for incident AD and all-cause dementia.

## 2. Methods

### 2.1 Study population

The National Health and Nutrition Examination Survey (NHANES) is a nationally-representative survey program of the health and nutritional status of the non-institutionalized US population operated by the National Center for Health Statistics (NCHS).^9^ This analysis used data from NHANES-III (1988-1994) and continuous NHANES (1999-2016), analyzed separately. We obtained exposure measures, key covariates, and sampling cluster, strata, and weights from NHANES and defined participants’ NHANES visit as our study baseline.

Medicare is the federal health insurance provider for individuals aged 65 years or older or with certain disabilities in the US. NCHS and the Centers for Medicare and Medicaid Services offer restricted-use data linkage between NHANES and Medicare records for participants who agreed to provide personal identification data.^10^ We used the Medicare Chronic Conditions Summary File (1999-2018) to identify cases of AD and all-cause dementia from comprehensive claims data. NHANES data were also linked to restricted-use data from the National Death Index (NDI) through 2019 to identify additional AD and all-cause dementia cases that were not identified in the Medicare claims.^11^ This analysis of existing data was approved by the University of Michigan Institutional Review Board (HUM00194918). All analyses were performed within a secure Federal Statistical Research Data Center operated by the US Census Bureau due to the restricted access to linked data. Further details of NHANES and Medicare data are available in the Supplement. The study design is presented in **Figure 1**.

**Figure 1.**
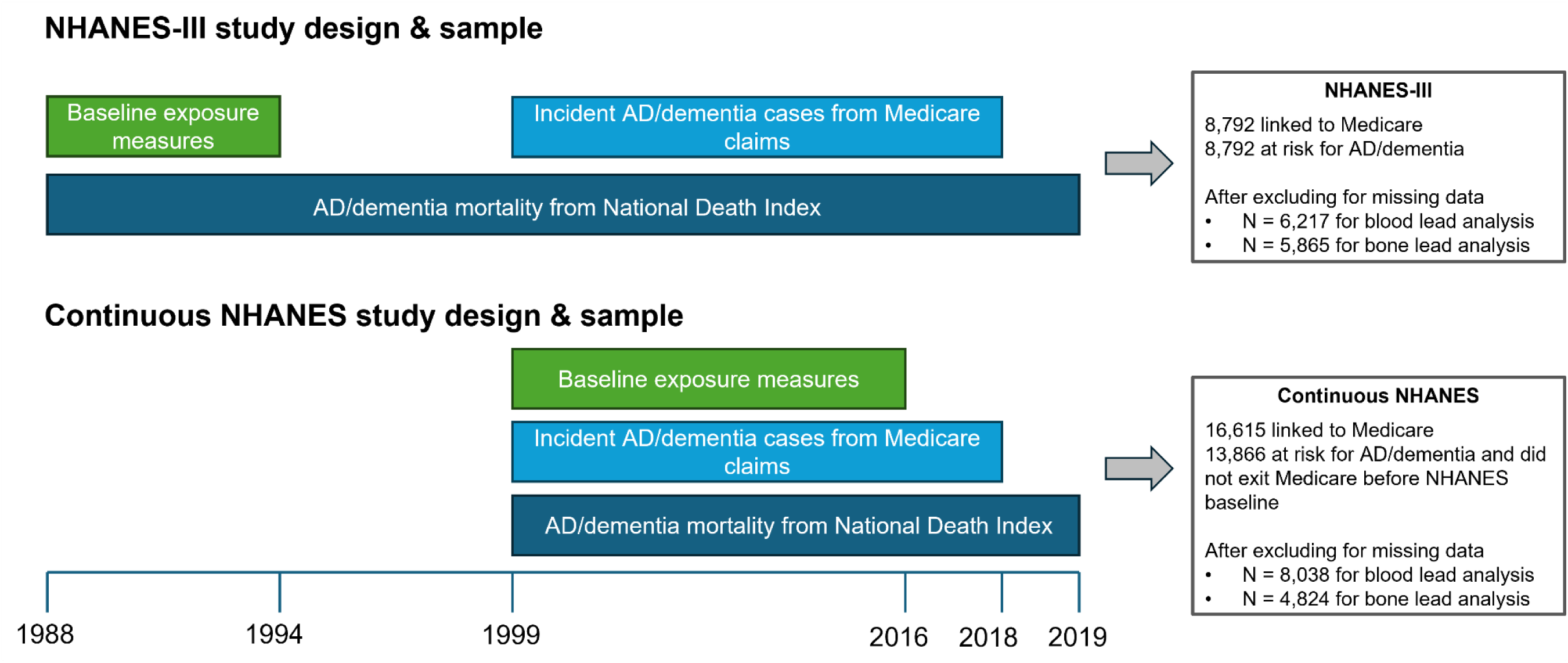
The study design flowchart with inclusion and exclusion criteria for the two independent samples. NHANES: National Health and Nutrition Examination Survey. AD: Alzheimer disease

### 2.2 Blood lead measurement

Whole blood lead concentrations (µg/dL) were measured at the Environmental Health Sciences Laboratory of the Center for Disease Control and Prevention’s National Center for Environmental Health. For NHANES-III and continuous NHANES 1999-2002 cycles, a simultaneous multielement atomic absorption spectrometer with Zeeman background correction was employed.^12,13^ Subsequent continuous NHANES cycles used inductively coupled plasma mass spectrometry.^14^ The limit of detection (LOD) for blood lead levels was 1 μg/dL for NHANES-III and ranged between 0.07-0.6 μg/dL for continuous NHANES, depending on cycle. NHANES imputed concentrations below LODs as LOD/√2. Complete laboratory protocol documentation is available on the NHANES website.^9^

### 2.3 Bone lead prediction

As direct bone lead measurements are unavailable in NHANES, we predicted concentrations (µg/g) of lead in patella and tibia for each participant using a machine learning-based ensemble of models comprised of eight algorithms (linear regression, generalized additive model, ridge regression, least absolute shrinkage and selection operator, elastic-net, classification and regression tree, random forest, and XGBoost).^15^ The models used seven predictors: blood lead concentration, age, education, occupation, body mass index (BMI), smoking status, and smoking pack-years. In a previous study, the Pearson correlation coefficients between observed and predicted values were 0.52 for tibia (cortical bone) lead and 0.58 for patella (trabecular bone) lead concentrations, indicating reasonable accuracy for evaluating the chronic health effects of cumulative lead exposure.^15^ The prediction models are publicly accessible via GitHub https://github.com/XinWangUmich/Bone-Lead-Prediction-Models). Estimated bone lead concentrations were unavailable for the continuous NHANES 2015-2016 cycle due to the absence of occupation information.

### 2.4 Outcome assessment

We defined incident cases as those with an AD or all-cause dementia diagnosis identified through either Medicare claims or mortality data. The dates of the first occurrence of AD and/or all-cause dementia were provided in the Medicare Chronic Conditions Summary File (1999-2018). AD is considered a specific subtype of dementia and AD cases were included in the all-cause dementia case sample. These conditions were pre-defined by NCHS according to the International Classification of Diseases, Ninth Revision (ICD-9), and Tenth Revision (ICD-10).^16^ The ICD-9 was valid until September 2015 with ICD-10 effective since October 2015. Diagnoses were based on the presence of at least one claim from inpatient services, skilled nursing facilities, home health agencies, hospital outpatient services, or carrier claims. The specific ICD-9 and ICD-10 codes defining AD and all-cause dementia diagnoses are detailed on the Medicare Chronic Conditions Data Warehouse website,^16^ with a summary in **Table S1**. Mortality data, including date of death and causes of death (primary and contributing) with ICD codes were sourced from the NDI’s Linked Mortality Restricted-use File (1988-2019). For consistency with Medicare data, we used the same ICD codes to identify AD and all-cause dementia mortality codes. Using the Medicare claims and NDI mortality data, we created separate variables to indicate AD and all-cause dementia outcome status. We calculated follow-up time as the years between NHANES participation and the first recorded date of the corresponding outcome event (AD/all-cause dementia) or the date of last follow-up (non-AD/non-dementia).

### 2.5 Covariate measures

Participant socio-demographic and behavioral data were collected during NHANES-III and continuous NHANES interviews and examinations. We included age (years), age^2^, sex (male/female), race/ethnicity (non-Hispanic White, non-Hispanic Black, Mexican American, other Hispanic (continuous NHANES only), and Other), educational attainment (less than high school, high school graduate, college and above), family income-to-poverty ratio (less than or equal to 1, greater than 1), smoking status (never, former, current), smoking pack-years, serum cotinine levels (ng/mL), alcohol consumption (0 drinks per day, 1 or more drinks per day), and BMI (kg/m^2^).

### 2.6 Statistical analysis

All participants included in this analysis were aged 65 or older by the end of 2018 (Medicare eligibility cutoff). We analyzed NHANES-III and continuous NHANES separately due to differences in measurements and sampling designs. Our analysis linked 8,792 NHANES-III participants with Medicare, all initially at risk for AD/all-cause dementia (i.e., participants without AD/all-cause dementia at baseline). After excluding participants with incomplete covariates or lead exposure, 6,217 participants remained for blood lead analysis and 5,865 for bone lead analysis. For the continuous NHANES sample, 16,615 participants were linked with Medicare. Of these, we excluded 2,729 participants who moved to Medicare Advantage or had Medicare claims for AD/all-cause dementia before their NHANES baseline examination, leaving 13,866 participants eligible at baseline. Exclusions for incomplete covariates or lead exposure resulted in final samples of 8,038 for blood lead analysis and 4,824 for bone lead analysis. The sample selection workflow is presented in **Figure S1**.

We accounted for the complex sampling design of NHANES by incorporating the survey cluster, strata, and sampling weights in all analyses, ensuring our findings are nationally-representative. We described continuous variables using mean and standard error and categorical variables using number and percentage. We compared the distributions of covariates among included and excluded participants with Medicare linkage. Within the included sample, we compared participants who did and did not develop AD, as well as participants who did and did not develop all-cause dementia. We compared groups using survey-weighted t-tests for continuous variables and survey-weighted Chi-square tests for categorical variables.

We employed survey-weighted Cox proportional hazards models to assess the associations between lead exposure measures and risks of AD and all-cause dementia in separate models. All models were adjusted for age, age^2^, sex, race/ethnicity, education, income-poverty ratio, smoking status, pack-years, serum cotinine, alcohol consumption, and BMI. Survival time for study participants was calculated from the lead measurement date to the occurrence of an AD or all-cause dementia event, or to death resulting from these conditions, depending on the data source of the outcome. Those without AD or all-cause dementia were right-censored at the date of their last available Medicare claim, death due to non-AD/non-dementia causes, or transfer to Medicare Advantage due to the inaccessibility of their claims data. Lead concentrations were log_2_-transformed to normalize their distributions and improve model goodness of fit. This approach allowed us to evaluate hazard ratios (HRs) per doubling of lead concentrations. To explore potential non-linear relationships, we also categorized lead concentrations into quartiles based on survey-weighted percentiles and calculated HRs for each of the upper three quartiles relative to the lowest quartile as the reference. Linear trends of the association across quartiles were tested by including quartiles as a continuous variable. Kaplan-Meier (K-M) curves were plotted to compare the adjusted survival probabilities of AD or all-cause dementia across different lead concentrations. For visualization, blood lead and estimated bone lead concentrations were dichotomized at the survey-weighted median. Joint inverse probability weighting was applied to account for potential confounding and sampling weight.

We calculated population attributable fractions (PAFs) by comparing the observed exposure distribution to a hypothetical reduced distribution, set at the 25^th^ survey-weighted percentile of each lead concentration.^17^ The PAF estimates the proportion of incident cases that could be prevented if population lead exposure were reduced to this level. The PAFs were derived from the survey-weighted Cox models with log_2_-transformed lead concentrations.

We conducted three sensitivity analyses. First, because dementia risk varies by sex,^18^ we added multiplicative interaction terms between lead concentrations and sex across all regression models to investigate whether sex modified the associations. Second, to discern if bone lead associations were independent of the most important predictors of estimated bone lead concentrations (age and blood lead),^15^ we additionally adjusted associations of estimated bone lead concentrations with AD and all-cause dementia for blood lead. Finally, we employed Fine-Gray competing risk regression to account for the competing risk of death in the estimation of AD and all-cause dementia risk. Deaths due to non-AD/non-dementia causes were categorized as a competing outcome. The competing risks models only incorporated sampling weights, not classes or strata, due to software limitations. All analyses were conducted using SAS (version 9.4, SAS Institute) and R (version 4.4.2), with statistical significance set at α < 0.05.

## 3. Results

### 3.1 Descriptive statistics

In continuous NHANES, 8,038 participants had a mean baseline age of 64.1 years (**Table 1**). During a mean follow-up period of 9.5 years, 1,260 participants developed all-cause dementia (186 dementia-related deaths), and 587 participants developed AD (154 AD-related deaths). Participants who developed AD/all-cause dementia had higher concentrations of blood lead, estimated patella lead, and estimated tibia lead, and were more likely to be female, non-smokers, and have lower education, income, alcohol consumption, and lower BMI. Participants excluded due to missing data tended to be older, have higher lead measurements, and have lower education levels (**Table S2)**.

**Table 1.** Baseline characteristics of 8,038 participants who had blood lead measured in continuous NHANES 1999-2016.

|  | **All**<br>**N = 8,038** | **Non-dementia**<br>**N = 6,778** | **All-cause**<br>**dementia**<br>**N = 1,260** | **P-value** | **Non-AD**<br>**N = 7,451** | **AD**<br>**N = 587** | **P-value** |
| --- | --- | --- | --- | --- | --- | --- | --- |
| **Continuous variables, mean (SE)** |  |  |  |  |  |  |  |
| Blood lead, µg/dL | 2.09 (0.03) | 2.05 (0.03) | 2.38 (0.07) | <.0001 | 2.08 (0.03) | 2.24 (0.07) | 0.04 |
| Estimated patella lead, µg/g<sup>a</sup> | 24.66 (0.16) | 23.87 (0.17) | 28.71 (0.31) | <.0001 | 24.32 (0.17) | 28.67 (0.39) | <.0001 |
| Estimated tibia lead, µg/g<sup>a</sup> | 16.65 (0.13) | 15.96 (0.14) | 20.18 (0.27) | <.0001 | 16.36 (0.13) | 20.12 (0.34) | <.0001 |
| Follow-up time, years | 9.46 (0.12) | 9.66 (0.13) | 7.98 (0.20) | <.0001 | 9.74 (0.13) | 8.53 (0.22) | <.0001 |
| Age, years | 64.13 (0.19) | 63.01 (0.20) | 72.18 (0.33) | <.0001 | 63.60 (0.19) | 73.21 (0.48) | <.0001 |
| Pack-years | 14.03 (0.37) | 14.02 (0.39) | 14.07 (0.98) | 0.96 | 14.12 (0.38) | 12.42 (1.32) | 0.21 |
| Serum cotinine, ng/mL | 45.38 (1.78) | 46.55 (1.93) | 37.34 (3.75) | 0.03 | 46.39 (1.86) | 29.13 (4.35) | 0.0003 |
| Body mass index, kg/m<sup>2</sup> | 28.78 (0.09) | 28.93 (0.10) | 27.74 (0.17) | <.0001 | 28.89 (0.10) | 27.07 (0.25) | <.0001 |
| **Categorical variables, n (%)** |  |  |  |  |  |  |  |
| Sex |  |  |  | <.0001 |  |  | <.0001 |
| Male | 3,822 (45.39%) | 3,267 (46.24%) | 555 (39.34%) |  | 3,588 (46.06%) | 234 (33.99%) |  |
| Female | 4,216 (54.61%) | 3,511 (53.76%) | 705 (60.66%) |  | 3,863 (53.94%) | 353 (66.01%) |  |
| Race/ethnicity |  |  |  | 0.01 |  |  | 0.02 |
| Non-Hispanic White | 4,618 (81.71%) | 3,787 (81.37%) | 831 (84.14%) |  | 4,222 (81.48%) | 396 (85.65%) |  |
| Non-Hispanic Black | 1,403 (7.51%) | 1,199 (7.41%) | 204 (8.19%) |  | 1,323 (7.54%) | 80 (7.02%) |  |
| Mexican American | 1,168 (3.40%) | 1,020 (3.50%) | 148 (2.67%) |  | 1,092 (3.43%) | 76 (2.87%) |  |
| Other Hispanic | 488 (3.35%) | 444 (3.51%) | 44 (2.14%) |  | 469 (3.45%) | 19 (1.62%) |  |
| Other | 361 (4.04%) | 328 (4.20%) | 33 (2.85%) |  | 345 (4.11%) | 16 (2.83%) |  |
| Education |  |  |  | <.0001 |  |  | <.0001 |
| Less than high school | 2,300 (17.10%) | 1,834 (15.71%) | 466 (27.07%) |  | 2,076 (16.45%) | 224 (28.06%) |  |
| High school graduate | 3,945 (53.43%) | 3,353 (53.44%) | 592 (53.34%) |  | 3,675 (53.44%) | 270 (53.25%) |  |
| College and above | 1,793 (29.47%) | 1,591 (30.85%) | 202 (19.60%) |  | 1,700 (30.11%) | 93 (18.69%) |  |
| Income-poverty ratio |  |  |  | <.0001 |  |  | 0.0004 |
| ≤1 | 1,191 (8.32%) | 965 (7.68%) | 226 (12.93%) |  | 1,092 (8.07%) | 99 (12.58%) |  |
| >1 | 6,847 (91.68%) | 5,813 (92.32%) | 1,034 (87.07%) |  | 6,359 (91.93%) | 488 (87.42%) |  |
| Smoking status |  |  |  | 0.02 |  |  | 0.01 |
| Never | 4,087 (50.13%) | 3,406 (49.61%) | 681 (53.85%) |  | 3,756 (49.77%) | 331 (56.21%) |  |
| Former | 2,739 (34.75%) | 2,308 (34.80%) | 431 (34.40%) |  | 2,531 (34.77%) | 208 (34.52%) |  |
| Current | 1,212 (15.12%) | 1,064 (15.59%) | 148 (11.74%) |  | 1,164 (15.46%) | 48 (9.28%) |  |
| Alcohol consumption |  |  |  | 0.01 |  |  | 0.002 |
| 0 drink/day | 7,023 (85.78%) | 5,886 (85.23%) | 1,137 (89.58%) |  | 6,483 (85.41%) | 540 (91.80%) |  |
| ≥1 drink/day | 1,015 (14.22%) | 892 (14.77%) | 123 (10.42%) |  | 968 (14.59%) | 47 (8.20%) |  |

SE: standard error; BMI: body mass index; AD: Alzheimer disease. NHANES: National Health and Nutrition Examination Survey
Survey weighted means and SEs were calculated for continuous variables. Unweighted frequencies and survey weighted percentages were calculated for categorical variables. Survey weighted t-tests were used to test continuous variables, and survey weighted Chi-square tests were used for categorical variables.
<sup>a</sup> Estimated patella and tibia lead were available in a subpopulation of 4,824 participants (3,902 non-dementia, 922 all-cause dementia, 4,381 non-AD, 443 AD) in the 1999-2014 cycles only.

In NHANES-III, 6,217 participants had a mean baseline age of 53.9 years (**Table S3**). Over a mean follow-up of 20.3 years, 1,543 participants developed all-cause dementia (235 dementia-related deaths) and 757 developed AD (188 AD-related deaths). Similar patterns of lead exposure and covariates by dementia status were observed as in the continuous NHANES sample, as well as between included and excluded participants (**Table S4**).

### 3.2 Associations between lead concentrations and incident AD

The K-M curves showed that participants with estimated bone lead concentrations above the medians experienced lower survival to AD during the follow-up, compared to those with bone lead concentrations below the medians (**Figure 2, Figure S2**).

**Figure 2.**
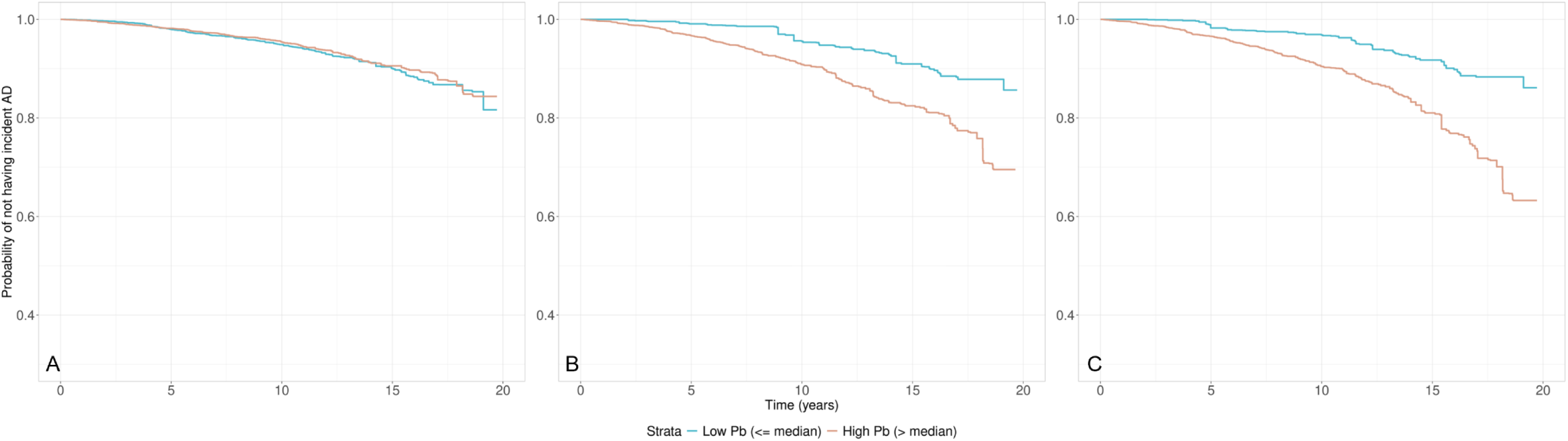
Kaplan-Meier (K-M) curves showing the probability of remaining free from Alzheimer disease (AD), stratified by median concentrations of (A) blood lead (1.7 µg/dL), (B) estimated patella lead (23.9 µg/g), and (C) estimated tibia lead (16.4 µg/g), in continuous NHANES. The curves account for survey weights and inverse probability weighting based on key covariates including age, age^2^, sex, race/ethnicity, education, income-poverty ratio, smoking status, pack-year, serum cotinine, alcohol consumption, and body mass index.

Multivariable adjusted survey-weighted Cox regression proportional hazards models showed higher estimated bone lead concentrations were associated with incident AD. In continuous NHANES, each doubling of estimated patella lead was associated with 1.74 (95% CI: 1.00-3.01, P=0.03) times higher hazard for incident AD, demonstrating a log-linear association between estimated patella lead and AD (**Table 2**). The highest estimated patella lead quartile had 2.96 (95% CI: 1.37-6.39, P for trend=0.11) times higher hazard for incident AD than the lowest quartile.

**Table 2.** Associations between lead and incident AD and all-cause dementia in continuous NHANES 1999-2016.

|  | Quartiles of lead concentrations<sup>a</sup> |  |  |  |  | Continuous lead concentrations |  |
| --- | --- | --- | --- | --- | --- | --- | --- |
|  | Quartile 1 | Quartile 2 | Quartile 3 | Quartile 4 | P for trend | Per doubling | P |
| **Association between lead and AD** |  |  |  |  |  |  |  |
| Blood lead, N = 8,038 |  |  |  |  |  |  |  |
| Range (µg/dL) | 0.18-1.19 | 1.20-1.69 | 1.70-2.49 | 2.50-54.00 |  |  |  |
| #Cases/Participants | 101/1,706 | 127/1,792 | 152/2,075 | 207/2,465 |  |  |  |
| HR (95% CI) | Ref | 1.04 (0.78, 1.38) | 0.80 (0.60, 1.07) | 0.84 (0.63, 1.12) | 0.16 | 0.90 (0.80, 1.02) | 0.14 |
| Estimated patella lead<sup>b</sup>, N = 4,824 |  |  |  |  |  |  |  |
| Range (µg/g) | 10.95-19.51 | 19.51-23.89 | 23.89-28.96 | 28.96-67.46 |  |  |  |
| #Cases/Participants | 14/857 | 54/1,151 | 137/1,271 | 238/1,545 |  |  |  |
| HR (95% CI) | Ref | 2.42 (1.31, 4.50) | 2.77 (1.40, 5.47) | 2.96 (1.37, 6.39) | 0.11 | 1.74 (1.00, 3.01) | 0.03 |
| Estimated tibia lead<sup>b</sup>, N = 4,824 |  |  |  |  |  |  |  |
| Range (µg/g) | 2.45-12.58 | 12.59-16.35 | 16.35-20.25 | 20.26-46.50 |  |  |  |
| #Cases/Participants | 13/809 | 55/1,114 | 126/1,287 | 249/1,614 |  |  |  |
| HR (95% CI) | Ref | 1.25 (0.69, 2.24) | 1.19 (0.62, 2.28) | 1.06 (0.51, 2.23) | 0.71 | 1.03 (0.60, 1.78) | 0.91 |
| **Association between lead and all-cause dementia** |  |  |  |  |  |  |  |
| Blood lead, N = 8,038 |  |  |  |  |  |  |  |
| Range (µg/dL) | 0.18-1.19 | 1.20-1.69 | 1.70-2.49 | 2.50-54.00 |  |  |  |
| #Cases/Participants | 204/1,706 | 239/1,792 | 330/2,075 | 487/2,465 |  |  |  |
| HR (95% CI) | Ref | 0.89 (0.73, 1.09) | 0.86 (0.71, 1.05) | 0.96 (0.79, 1.16) | 0.84 | 0.99 (0.92, 1.08) | 0.91 |
| Estimated patella lead<sup>b</sup>, N = 4,824 |  |  |  |  |  |  |  |
| Range (µg/g) | 10.95-19.51 | 19.51-23.89 | 23.89-28.96 | 28.96-67.46 |  |  |  |
| #Cases/Participants | 40/857 | 121/1,151 | 257/1,271 | 504/1,545 |  |  |  |
| HR (95% CI) | Ref | 1.48 (1.02, 2.16) | 1.66 (1.09, 2.53) | 2.15 (1.33, 3.46) | 0.01 | 1.82 (1.26, 2.63) | 0.004 |
| Estimated tibia lead<sup>b</sup>, N = 4,824 |  |  |  |  |  |  |  |
| Range (µg/g) | 2.45-12.58 | 12.59-16.35 | 16.35-20.25 | 20.26-46.50 |  |  |  |
| No. Cases | 34/809 | 121/1,114 | 245/1,287 | 522/1,614 |  |  |  |
| HR (95% CI) | Ref | 1.22 (0.83, 1.79) | 1.28 (0.83, 1.97) | 1.41 (0.86, 2.32) | 0.27 | 1.63 (1.14, 2.34) | 0.01 |

AD: Alzheimer disease. All models were adjusted for age, age<sup>2</sup>, sex, race/ethnicity, education, income-poverty ratio, smoking status, pack-year, serum cotinine, alcohol consumption, and body mass index.
<sup>a</sup> Quartiles of lead concentrations were based on survey-weighted distributions.
<sup>b</sup> Bone lead was available in NHANES 1999-2014.

**Table 3.** Population attributable fractions for AD and all-cause dementia in continuous NHANES 1999-2016.

| **PAF (95% CI)** | **Blood lead** | **Estimated patella lead** | **Estimated tibia lead** |
| --- | --- | --- | --- |
| Reference concentrations (25<sup>th</sup> percentile) | 1.2 µg/dL | 19.5 µg/g | 12.6 µg/g |
| **AD** | -4% (-16%, 7%) | 16% (-3%, 34%) | 1% (-28%, 26%) |
| **All-cause dementia** | -0.4% (-10%, 9%) | 18% (2%, 32%) | 17% (-2%, 33%) |

PAF: population attributable fraction; AD: Alzheimer disease. PAFs were derived from survey-weighted Cox proportional hazards models utilizing log<sub>2</sub>-transformed lead concentrations. Calculations compared observed exposure distributions to the 25<sup>th</sup> percentile of each lead concentration as the theoretical low risk exposure distributions.

In NHANES-III, no associations between lead concentrations and incident AD were observed (**Table S5**).

### 3.3 Associations between lead concentrations and incident all-cause dementia

Similar to the results for AD, the K-M curves demonstrated that participants with estimated bone lead concentrations above the medians had lower survival rates for all-cause dementia during the follow-up period compared to those with estimated bone lead concentrations below the medians (**Figure S3, Figure S4**).

Adjusted survey-weighted Cox regression models in continuous NHANES showed higher concentrations of both estimated patella and tibia lead were associated with incident all-cause dementia (**Table 2**). The highest quartile for estimated patella lead was associated with 2.15 (95% CI: 1.33-3.46, P for trend=0.01) times higher hazard for incident all-cause dementia, relative to the lowest quartile. When modeling estimated lead levels continuously, a positive log-linear association between estimated patella lead and all-cause dementia was also observed (HR: 1.82, 95% CI: 1.26-2.63, P=0.004). Similar relationships between estimated tibia lead (quartiles and continuous) and incident all-cause dementia were identified (**Table 2**).

NHANES-III analyses additionally showed higher estimated bone lead was associated with incident all-cause dementia (**Table S5**). The highest quartile of estimated tibia lead was associated with 1.87 (95% CI: 1.10-3.19, P for trend = 0.44) times higher hazard of incident all-cause dementia. A similar relationship between the highest estimated patella lead quartile and incident all-cause dementia was also observed (HR: 1.67, 95% CI: 1.09-2.56).

### 3.4 Population attributable fractions (PAF)

In continuous NHANES, if estimated patella lead concentrations dropped to the 25^th^ percentile (19.5 µg/g), the PAF for incident all-cause dementia was 18% (95% CI: 2%-32%).

In NHANES-III, no PAFs were statistically significant (**Table S6**).

### 3.5 Sensitivity analyses

No significant effect modification by sex was found in either sample (**Table S7**, **Table S8**). In bone lead models with additional adjustment for blood lead, similar associations between estimated patella and tibia lead concentrations and AD/all-cause dementia were observed (**Table S9, Table S10**).

In competing risk analyses, the associations between estimated patella lead and AD/all-cause dementia in continuous NHANES were attenuated (**Table S11**). However, we also observed a significant sub-hazard ratio (SHR) for the association between blood lead and all-cause dementia (SHR: 1.09, 95% CI: 1.01-1.17) (**Table S11**). Lastly, in both competing risks and mortality-specific Cox models, lead exposures were consistently associated with increased hazard of mortality in NHANES-III (**Table S12**) and in some continuous NHANES bone lead analyses (**Table S11**).

## 4. Discussion

We conducted a comprehensive analysis of associations between lead exposure and incident AD and all-cause dementia using data from over 14,000 participants in NHANES linked with Medicare claims for follow-up periods of up to 30 years. We found that higher estimated bone (patella and tibia) lead concentrations at baseline were significantly associated with increased hazard of AD and all-cause dementia, suggesting that cumulative lead exposure is an important environmental risk factor for these neurodegenerative conditions. The Lancet commission has identified numerous modifiable factors for preventing dementia including air pollution,^19^ and this study contributes to a body of evidence that lead exposure may be another factor. This study marks an important step towards expanding the scope of exposures investigated with dementia towards the goal of investigating the totality of exposures, broadly termed the exposome.^20^

Our findings suggest that approximately 18% of new all-cause dementia cases in the US each year could be linked to cumulative lead exposure. Lead absorbed from environmental sources over a lifetime can accumulate in various bone sites, especially during periods of mineral deposition, where it remains for years to decades.^8^ Lead is later released during bone resorption, making bone lead the primary source of ongoing internal lead exposure in aging populations, despite recent reductions in environmental lead levels. Our results highlight a potentially significant, and overlooked, contribution of cumulative lead exposure-especially from bone lead stores-to the risk of AD and all-cause dementia in the US.

Our study builds on previous epidemiologic analyses of lead exposure and AD. A systematic review by Brown et al. of case-control studies comparing lead concentrations in blood, cerebrospinal fluid, hair/nail, and urine in AD patients versus controls found insufficient evidence to establish a link between lead and AD, emphasizing the need for longitudinal studies with bone lead concentrations to evaluate the effects of chronic lead exposure.^21^ Horton et al. investigated relationships between blood lead concentrations and AD mortality in a cohort from NHANES 1999-2008 and public-use linked mortality files until 2014.^22^ They found an association, though not statistically significant, between higher blood lead concentration and increased AD mortality. Our study expands on these findings, using a larger prospective cohort with estimated bone lead concentrations and AD/all-cause dementia incidence data from both mortality files and Medicare claims, strengthening the evidence for a potential association between lead exposure and AD. A recent study estimating lead exposure from residential information showed lead from industrial releases may be linked to lower cognition in older adults.^23^ Lead exposures are linked to other significant, age-related brain disorders, notably stroke and Parkinson disease.^24,25^ These established relationships are broadly consistent with our findings of associations between higher lead exposure and increased AD/all-cause dementia incidence, suggesting a broad impact of lead exposures on age-related neurologic disorders.

In our study, stronger associations were observed in continuous NHANES compared to NHANES-III. NHANES-III occurred from 1988-1994, while Medicare linkage includes claims dating back only to 1999. It is possible that a higher proportion of NHANES-III participants had already developed these conditions before 1999. Greater pre-enrollment prevalence in NHANES-III, linked to Medicare claims beginning in 1999, could result in an underestimation of the associations between lead and incident cases in NHANES-III. The longer follow-up time in NHANES-III may contribute to weaker observed associations due to time-varying hazard ratios and depletion of susceptible participants, where participants who were more susceptible to lead toxicity may have developed AD and all-cause dementia earlier were progressively excluded from the analysis over time.

The present study identified a significant association with estimated bone lead, but not with measured blood lead. This finding suggests that circulating lead, as reflected by blood lead, may not be a relevant indicator of long-term cumulative lead exposure or suitable for assessing risk for chronic diseases such as AD and all-cause dementia. Over the past several decades, blood lead levels have declined in the US, largely due to the phaseout of leaded gasoline and lead-based residential paint.^26^ Despite this decrease in blood lead, the total body burden, as indicated by bone lead, remains stable until bone turnover rates increase.^27^ Most adults included in the present study were born before 1980, a period of elevated environmental lead exposure, and they likely have substantial cumulative lead stores even if their current blood lead levels are relatively low. This observation implies that adults in the US who were born before 1980 may be at increased risk of neurodegenerative diseases due to their cumulative lead burden.

It is important note that analyses using hemoglobin-corrected blood lead levels, which have been proposed to account for potential confounding by anemia, produced nearly identical results (data not shown). The association between whole blood lead levels and dementia risk may be confounded by anemia, since anemia can influence blood lead levels (most lead in the blood is bound to red blood cells).^27^ Anemia and iron deficiency have also been linked to higher dementia risk.^28,29^

Lead accumulation is implicated in AD pathology. Research in preclinical models showed that lead exposure upregulates expression of the amyloid precursor protein and increases enzymatic activity of β-secretase, resulting in higher production of the neurotoxic beta-amyloid (Aβ)42 peptide, increased Aβ deposition, and promotion of brain plaque proliferation.^30,31^ Lead may disrupt clearance of Aβ by impairing pathways mediated by the lipoprotein receptor-related protein-1 and insulin-degrading enzyme, affecting glial cell functions, and altering blood-brain barrier integrity.^32,33^ Lead exposure may also contribute to formation of neurofibrillary tangles by enhancing production and phosphorylation of tau protein and impairing structural stability of microtubules in neurons,^4^ further exacerbating AD-related neurodegeneration. Toxicological impairments and pathologies related to other potentially relevant pathways are observed with lead exposures. Lead can induce oxidative stress and neuroinflammation,^5^ impair brain endothelial function and cerebral blood vessels,^34^ and is associated with cerebrovascular dementia risk factors such as peripheral arterial disease and atherosclerosis.^35,36^ Lead exposure may induce aggregation and inclusion of TDP-43, a major pathological protein in frontotemporal dementia and amyotrophic lateral sclerosis.^37^ Lead is suggested to contribute to dementia with Lewy bodies and Parkinson’s disease by disrupting the dopaminergic system, as evidenced by changes in dopamine release, neuron activity, dependent behaviors, synthesis, turnover, and uptake in certain brain areas.^38^

The public health implications of our study are underscored by the relatively large PAF of reducing lead burden on AD/all-cause dementia incidence. These predicted effects are significantly larger than predicted effects of reducing established modifiable risk factors in the Lancet Commission on dementia.^19^ It should be noted that the PAFs reported here do not account for communalities, i.e., clustering of AD and all-cause dementia risk factors such as lower levels of education, hearing loss, high low-density lipoprotein cholesterol, depression, and hypertension.^19^ Therefore, PAFs corrected for these communalities in cumulative lead exposure may be smaller. However, given the widespread presence of lead and its well-established public health impacts, our study supports the need for policy-making and preventive strategies that specifically target lead exposure reduction, potentially lowering AD and all-cause dementia risks and alleviating its burden on healthcare systems.

Our study has several limitations to consider. The accuracy of AD and all-cause dementia ascertainment using Medicare claims may not be perfect. Previous research indicates that, compared to clinical dementia assessments from the Aging Demographics and Memory Study cohort of the Health and Retirement Study (the “gold standard”), Medicare claims data had a sensitivity and specificity of 85% and 89% for dementia, and 64% and 95% for AD, respectively.^39^ Medicare claims are a cost-effective and reasonably accurate method for large-scale population-based studies. Our study only captured AD and all-cause dementia cases in individuals aged 65 and older, excluding those who developed these conditions before becoming eligible for Medicare. Given the relatively low incidence rates of early-onset AD and all-cause dementia, this limitation likely had minimal impact. Most importantly, our estimates of bone lead concentrations were derived from predictive models rather than direct measurements. While direct measurement using K-shell X-ray fluorescence is the gold standard for assessing bone lead,^7^ it is impractical for large population-based studies due to technical and financial constraints. These predictive models, based on blood lead concentrations and routinely collected variables, applied through machine-learning algorithms, offer a viable alternative for assessing the health effects of cumulative lead exposure when direct bone lead measurements are unavailable. However, the moderate predictive performance of these models (correlation coefficients of 0.52 for tibia lead and 0.58 for patella lead) raises the possibility that the observed associations may have been underestimated.

Our study observes that higher cumulative lead exposure is associated with increased hazard of incident AD and all-cause dementia, leveraging data from over 14,000 participants in a long-term prospective cohort representative of the US general population. These findings underscore the role of lead as an environmental risk factor for these neurodegenerative conditions and advocate for urgent public health interventions to mitigate lead exposure, potentially reducing the burden of AD and dementia. The study also emphasizes the necessity of incorporating environmental risk factors into dementia research and clinical practice to better understand and mitigate their contribution to the development of neurodegenerative diseases.

## Research in Context

### Evidence before this study

Before initiating this study, we conducted a comprehensive literature search using multiple databases including PubMed and Web of Science. We searched for studies published from January 1980 to December 2024, without language restrictions, using search terms such as “lead exposure,” “bone lead,” “patella lead”, “tibia lead”, “blood lead,” “Alzheimer disease,” “dementia,” “cognitive decline,” and related terms. Reference lists of relevant journal articles and book chapters were also reviewed to ensure thorough coverage. Studies were included if they evaluated the association between lead exposure and dementia related outcomes. The quality of the available evidence was assessed using standard risk-of-bias criteria. Several cross-sectional and case-control studies indicated a potential link between lead exposure and dementia, and a few prospective studies provided mixed results, highlighting the need for a large, nationally representative prospective cohort analysis.

### Added value of this study

Our study adds significant value by leveraging data from over 14,000 participants in the NHANES cohorts linked with Medicare claims and mortality records, offering up to 30 years of follow-up. Unlike previous studies that primarily focused on blood lead concentrations, we used both measured blood lead and machine-learning–predicted bone lead concentrations (patella and tibia) to assess cumulative lead exposure. This approach allowed us to demonstrate the associations between higher estimated bone lead and increased risks of incident AD and all-cause dementia. Additionally, our study estimated population attributable fractions, suggesting that a substantial proportion of AD/all-cause dementia cases could potentially be prevented if lead exposure were reduced.

### Implications of all the available evidence

Collectively, the existing literature combined with our new findings imply that cumulative lead exposure is an environmental risk factor for AD and dementia. These results underscore the urgency for public health policies aimed at reducing lead exposure in the population. Furthermore, the evidence advocates for the integration of environmental risk assessments into clinical practice and dementia research. Future studies should continue to refine exposure assessment methods and explore the biological mechanisms linking lead exposure to neurodegeneration to better inform targeted prevention strategies.

## Acknowledgements

We thank Dieudonne Nahigombeye at the Centers for Disease Control and Prevention for his assistance with accessing restricted data and reviewing the analysis outputs. This research was supported by the National Institute on Aging (R01 AG070897, K01 AG084821, P30 AG072931) and the National Institute of Environmental Health Sciences (P30 ES017885).

## Data Sharing Statement

NHANES datasets are publicly available at https://www.n.cdc.gov/nchs/nhanes. Bone lead prediction models are publicly available at https://github.com/XinWangUmich/Bone-Lead-Prediction-Models. Restricted-use linked Medicare and National Death Index datasets must be analyzed in Federal Statistical Research Data Centers and require an application process to access.

