## Supplementary materials for "Exposure to Lead and Incidence of Alzheimer Disease and All-Cause Dementia in the United States"

**Supplementary Appendix**

This document contains Supplementary Methods, 12 supplemental tables, and 4 supplemental figures.

**Supplementary Methods**

**Study Population:** Baseline data for this study were from the National Health and Nutrition Examination Survey (NHANES), a program initiated in the 1960s by the National Center for Health Statistics (NCHS) to assess the health and nutritional status of the non-institutionalized US population.^1^ NHANES-III (conducted 1988-1994) expanded the scope of the initial surveys, including collecting data on environmental exposures. From 1999 onwards, NHANES became a continuous program, with data released biennially. Each cycle constitutes an independent sample from a multi-stage complex sampling design and includes interviews, examinations, and laboratory tests for comprehensive health, nutritional, and environmental data collection. This study used both NHANES-III and continuous NHANES (1999-2016). All procedures were approved by the NCHS Ethics Review Board, with participant consent obtained in writing.

Medicare is the federal health insurance provider for individuals aged 65 years or older or with certain disabilities in the US. NHANES participants were linked to Medicare claims by NCHS, and our study team applied and received approval for the restricted-used data access.^2^ We used the Medicare Chronic Conditions Summary File (1999-2018) to identify cases of Alzheimer disease (AD) and all-cause dementia based on the definitions created by the Centers for Medicare and Medicaid Services. The all-cause dementia outcome in our analysis was based on the Medicare chronic condition definition for Alzheimer disease and related dementias (ADRD); notably, ADRD does not include the ICD codes for dementia with Lewy bodies. NHANES data were also linked to death certificate records from the National Death Index up to 2019,^3^ which was used to identify additional AD and all-cause dementia cases that were not identified in the Medicare claims. We evaluated AD and all-cause dementia in separate analyses. Because AD is a specific subtype of dementia, AD cases were also considered in the all-cause dementia outcome. If a participant had claims for both AD and a separate non-AD dementia, we considered the diagnosis event that occurred first for the all-cause dementia analysis.

In the present analysis, participants were included if they were 65 years or older by the end of available Medicare follow-up in 2018. Participants’ age in 2018 was estimated by calculating the number of years between NHANES participation and 2018, then adding it to their age at the time of the NHANES examination. For example, a participant aged 40 years old in NHANES-III that was examined in 1990 would be 68 years in 2018 and therefore eligible for inclusion. This allows for a delayed-entry study design, where participant inclusion in the study was staggered throughout the follow-up period based on the time of NHANES data collection and Medicare eligibility.

**Supplementary Tables**

**Table S1.** International Classification of Disease (ICD) codes used to define Alzheimer disease (AD) and all-cause dementia through Medicare claims and through the National Death Index. These classifications were provided through the Medicare Chronic Conditions Summary File. Details of the chronic condition including classification algorithms can be found at the Chronic Condition Data Warehouse at <https://www2.ccwdata.org/web/guest/condition-categories-chronic#cc27>.

|  | **ICD-9** | **ICD-10** |
| --- | --- | --- |
| **AD** | **331.0:** Alzheimer’s disease | **G30.0:** Alzheimer’s disease with early onset  **G30.1:** Alzheimer’s disease with late onset  **G30.8:** Other Alzheimer’s disease  **G30.9:** Alzheimer’s disease, unspecified |
| **All-cause dementia** | **331.0:** Alzheimer’s disease  **331.11:** Pick’s disease (frontotemporal dementia)  **331.19:** Other frontotemporal dementia  **331.2:** Senile degeneration of brain  **331.7:** Cerebral degeneration in diseases classified elsewhere  **290.0:** Senile dementia, uncomplicated  **290.10:** Presenile dementia, uncomplicated  **290.11:** Presenile dementia with delirium  **290.12:** Presenile dementia with delusional features  **290.13:** Presenile dementia with depressive features  **290.20:** Senile dementia with delusional features  **290.21:** Senile dementia with depressive features  **290.3:** Senile dementia with delirium  **290.40:** Vascular dementia, uncomplicated  **290.41:** Vascular dementia with delirium  **290.42:** Vascular dementia with delusions  **290.43:** Vascular dementia with depressed mood  **294.0:** Amnestic disorder in conditions classified elsewhere  **294.10:** Dementia in conditions classified elsewhere without behavioral disturbance  **294.11:** Dementia in conditions classified elsewhere with behavioral disturbance  **294.20:** Dementia, unspecified, without behavioral disturbance  **294.21:** Dementia unspecified, with behavioral disturbance  **294.8:** Other persistent mental disorders due to conditions classified elsewhere  **797:** Senility without mention of psychosis | **F01.50:** Vascular dementia, unspecified severity, without behavioral disturbance, psychotic disturbance, mood disturbance, and anxiety  **F01.51:** Vascular dementia with behavioral disturbance  **F02.80:** Dementia in other diseases classified elsewhere, unspecified severity, without behavioral disturbance, psychotic disturbance, mood disturbance, and anxiety  **F02.81:** Dementia in other diseases classified elsewhere with behavioral disturbance  **F03.90:** Unspecified dementia, unspecified severity, without behavioral disturbance, psychotic disturbance, mood disturbance, and anxiety  **F03.91:** Unspecified dementia with behavioral disturbance  **F04:** Amnestic disorder due to known physiological condition  **F05:** Delirium due to known physiological condition  **F06.1:** Catatonic disorder due to known physiological condition  **F06.8:** Other specified mental disorders due to known physiological condition  **G13.8:** Systemic atrophy primarily affecting central nervous system in other diseases classified elsewhere  **G30.0:** Alzheimer’s disease with early onset  **G30.1:** Alzheimer’s disease with late onset  **G30.8:** Other Alzheimer’s disease  **G30.9:** Alzheimer’s disease, unspecified **G31.01:** Pick’s disease (frontotemporal dementia)  **G31.09:** Other frontotemporal neurocognitive disorder  **G31.1:** Senile degeneration of brain, not elsewhere classified  **G31.2:** Degeneration of nervous system due to alcohol  **G94:** Other disorders of brain in diseases classified elsewhere  **R41.81:** Age-related cognitive decline  **R54:** Age-related physical debility |

Cases were identified based on the presence of at least one claim from inpatient services, skilled nursing facilities, home health agencies, hospital outpatient services, or carrier claims with a qualifying ICD code. The ICD-9 was valid until September 2015, and ICD-10 has been effective since October 2015.

**Table S2.** Characteristics of included versus excluded continuous NHANES participants with Medicare claims linkage (N = 13,866)

|  | **All**  **N = 13,866** | **Included**  **N = 8,038** | **Excluded**  **N = 5,828** | **P-value** |
| --- | --- | --- | --- | --- |
| **Continuous variables, mean (SE)** | | | | |
| Blood lead, ug/dL | 2.13 (0.03) | 2.09 (0.03) | 2.24 (0.05) | 0.003 |
| Missing | 2,211 | 0 | 2,211 |  |
| Estimated patella lead, µg/g^a^ | 25.32 (0.15) | 24.66 (0.16) | 27.18 (0.25) | <.0001 |
| Missing | 6,955 | 3,214 | 3,741 |  |
| Estimated tibia lead, µg/g^a^ | 17.15 (0.12) | 16.65 (0.13) | 18.54 (0.21) | <.0001 |
| Missing | 6,955 | 3,214 | 3,741 |  |
| Follow-up time, years | 8.63 (0.11) | 9.46 (0.12) | 7.24 (0.16) | <.0001 |
| Age, years | 65.29 (0.16) | 64.13 (0.19) | 67.23 (0.20) | <.0001 |
| Pack-years | 15.50 (0.32) | 14.03 (0.37) | 18.12 (0.54) | <.0001 |
| Missing | 352 | 0 | 352 |  |
| Serum cotinine, ng/mL | 47.14 (1.50) | 45.38 (1.78) | 50.92 (2.38) | 0.0514 |
| Missing | 1,418 | 0 | 1,418 |  |
| Body mass index, kg/m^2^ | 28.89 (0.09) | 29.10 (0.13) | 28.78 (0.09) | 0.0167 |
| Missing | 1,043 | 0 | 1,043 |  |
| **Categorical variables, n (%)** | | | | |
| Sex |  |  |  | 0.0146 |
| Male | 6,911 (46.30%) | 3,822 (45.39%) | 3,089 (47.83%) |  |
| Female | 6,955 (53.70%) | 4,216 (52.17%) | 2,739 (52.17%) |  |
| Race/ethnicity |  |  |  | <.0001 |
| Non-Hispanic White | 7,543 (79.59%) | 4,618 (81.71%) | 2,925 (76.05%) |  |
| Non-Hispanic Black | 2,648 (8.55%) | 1,403 (7.51%) | 1,245 (10.30%) |  |
| Mexican American | 2,054 (3.70%) | 1,168 (3.40%) | 886 (4.20%) |  |
| Other Hispanic | 887 (3.44%) | 488 (3.35%) | 399 (3.59%) |  |
| Other | 734 (4.72%) | 361 (4.04%) | 373 (5.85%) |  |
| Education |  |  |  | <.0001 |
| Less than high school | 4,566 (20.27%) | 2,300 (17.10%) | 2,266 (25.60%) |  |
| High school graduate | 6,551 (52.63%) | 3,945 (53.43%) | 2,606 (51.27%) |  |
| College and above | 2,723 (27.10%) | 1,793 (29.47%) | 930 (23.12%) |  |
| Missing | 26 | 0 | 26 |  |
| Income-poverty ratio |  |  |  | <.0001 |
| <=1 | 2,150 (9.88%) | 1,191 (8.32%) | 959 (13.07%) |  |
| >1 | 10,586 (90.12%) | 6,847 (91.68%) | 3,739 (86.93%) |  |
| Missing | 1,130 | 0 | 1,130 |  |
| Smoking status |  |  |  | <.0001 |
| Never | 6,528 (46.88%) | 4,087 (50.13%) | 2,441 (41.44%) |  |
| Former | 5,203 (37.87%) | 2,739 (34.75%) | 2,464 (43.09%) |  |
| Current | 2,120 (15.25%) | 1,212 (15.12%) | 908 (15.47%) |  |
| Missing | 15 | 0 | 15 |  |
| Alcohol consumption |  |  |  | 0.0863 |
| 0 drink/day | 9,033 (86.21%) | 7,023 (85.78%) | 2,010 (87.88%) |  |
| ≥1 drink/day | 1,246 (13.79%) | 1,015 (14.22%) | 231 (12.12%) |  |
| Missing | 3,587 | 0 | 3,587 |  |

Note: Survey-weighted means and standard errors were calculated for continuous variables. Unweighted frequencies and survey-weighted percentages were calculated for categorical variables. Survey-weighted t-tests were used to test continuous variables, and survey-weighted Chi-square tests were used for categorical variables.

**Table S3.** Baseline characteristics of 6,217 participants who had blood lead measured in NHANES-III.

|  | **All**  **N = 6,217** | **Non-dementia**  **N= 4,674** | **All-cause dementia**  **N = 1,543** | **P-value** | **Non-AD**  **N = 5,460** | **AD**  **N = 757** | **P-value** |
| --- | --- | --- | --- | --- | --- | --- | --- |
| **Continuous variables, mean (SE)** | | | | | | | |
| Blood lead, µg/dL | 3.83 (0.10) | 3.80 (0.11) | 3.96 (0.10) | 0.10 | 3.83 (0.10) | 3.86 (0.13) | 0.84 |
| Estimated patella lead, µg/g^a^ | 21.67 (0.25) | 20.35 (0.25) | 27.47 (0.30) | <.0001 | 21.11 (0.26) | 27.36 (0.40) | <.0001 |
| Estimated tibia lead, µg/g^a^ | 12.95 (0.24) | 11.62 (0.24) | 18.78 (0.25) | <.0001 | 12.37 (0.25) | 18.76 (0.35) | <.0001 |
| Follow-up time, years | 20.33 (0.26) | 21.05 (0.27) | 17.13 (0.28) | <.0001 | 20.83 (0.27) | 17.44 (0.38) | <.0001 |
| Age, years | 53.93 (0.48) | 51.51 (0.46) | 64.63 (0.46) | <.0001 | 52.81 (0.48) | 65.00 (0.51) | <.0001 |
| Pack-years | 14.26 (0.47) | 14.12 (0.54) | 14.85 (0.93) | 0.51 | 14.25 (0.48) | 14.35 (1.22) | 0.93 |
| Serum cotinine, ng/mL | 70.24 (2.61) | 74.05 (2.66) | 53.75 (5.59) | 0.0007 | 71.95 (2.70) | 53.65 (8.52) | 0.04 |
| Body mass index, kg/m^2^ | 27.36 (0.14) | 27.36 (0.16) | 27.36 (0.23) | 0.99 | 27.36 (0.14) | 27.40 (0.31) | 0.91 |
| **Categorical variables, n (%)** | | | | | | | |
| Sex |  |  |  | <.0001 |  |  | <.0001 |
| Male | 2,827 (46.74%) | 2,191 (48.62%) | 636 (38.42%) |  | 2,533 (47.90%) | 294 (35.31%) |  |
| Female | 3,390 (53.26%) | 2,483 (51.38%) | 907 (61.58%) |  | 2,927 (52.10%) | 463 (64.69%) |  |
| Race/ethnicity |  |  |  | 0.17 |  |  | 0.30 |
| Non-Hispanic White | 3,181 (82.59%) | 2,308 (82.38%) | 873 (83.51%) |  | 2,747 (82.48%) | 434 (83.63%) |  |
| Non-Hispanic Black | 1,416 (8.15%) | 1,085 (7.97%) | 331 (8.95%) |  | 1,254 (8.08%) | 162 (8.81%) |  |
| Mexican American | 1,376 (3.39%) | 1,081 (3.53%) | 295 (2.75%) |  | 1,240 (3.51%) | 136 (2.19%) |  |
| Other | 244 (5.87%) | 200 (6.12%) | 44 (4.79%) |  | 219 (5.92%) | 25 (5.37%) |  |
| Education |  |  |  | <.0001 |  |  | <.0001 |
| Less than high school | 2,545 (24.69%) | 1,733 (21.63%) | 812 (38.23%) |  | 2,153 (23.34%) | 392 (38.01%) |  |
| High school graduate | 2,779 (52.34%) | 2,209 (53.67%) | 570 (46.45%) |  | 2,490 (52.81%) | 289 (47.73%) |  |
| College and above | 893 (22.97%) | 732 (24.70%) | 161 (15.32%) |  | 817 (23.85%) | 76 (14.26%) |  |
| Income-poverty ratio |  |  |  | <.0001 |  |  | 0.0002 |
| <=1 | 1,104 (8.40%) | 767 (7.43%) | 337 (12.70%) |  | 941 (7.99%) | 163 (12.51%) |  |
| >1 | 5,113 (91.60%) | 3,907 (92.57%) | 1,206 (87.30%) |  | 4,519 (92.01%) | 594 (87.49%) |  |
| Smoking status |  |  |  | 0.0002 |  |  | 0.07 |
| Never | 2,953 (44.05%) | 2,159 (43.12%) | 794 (48.18%) |  | 2,556 (43.60%) | 397 (48.52%) |  |
| Former | 1,949 (33.35%) | 1,448 (32.90%) | 501 (35.32%) |  | 1,696 (33.23%) | 253 (34.51%) |  |
| Current | 1,315 (22.60%) | 1,067 (23.98%) | 248 (16.50%) |  | 1,208 (23.17%) | 107 (16.97%) |  |
| Alcohol consumption |  |  |  | 0.01 |  |  | 0.048 |
| 0 drink/day | 5,503 (87.12%) | 4,093 (86.52%) | 1,410 (89.74%) |  | 4,809 (86.78%) | 694 (90.42%) |  |
| ≥1 drink/day | 714 (12.88%) | 581 (13.48%) | 133 (10.26%) |  | 651 (13.22%) | 63 (9.58%) |  |

SE: standard error; BMI: body mass index; AD: Alzheimer disease.

Survey-weighted means and SEs were calculated for continuous variables. Unweighted frequencies and survey-weighted percentages were calculated for categorical variables. Survey-weighted t-tests were used to test continuous variables, and survey weighted Chi-square tests were used for categorical variables.

^a^ Estimated patella and tibia lead were available in a subpopulation of 5,865 participants (4,427 non-dementia, 1,438 all-cause dementia; 5,160 non-AD, 705 AD).

**Table S4.** Characteristics of included versus excluded NHANES-III participants with Medicare claims linkage (N = 8,792)

|  | **All**  **N = 8,792** | **Included**  **N = 6,217** | **Excluded**  **N = 2,575** | **P-value** |
| --- | --- | --- | --- | --- |
| **Continuous variables, mean (SE)** | | | | |
| Blood lead, ug/dL | 3.87 (0.10) | 3.83 (0.10) | 4.07 (0.14) | 0.0316 |
| Missing | 1,076 | 0 | 1,076 |  |
| Estimated patella lead, µg/g^a^ | 22.05 (0.26) | 21.67 (0.25) | 25.20 (0.58) | <.0001 |
| Missing | 1,888 | 352 | 1,536 |  |
| Estimated tibia lead, µg/g^a^ | 13.29 (0.24) | 12.95 (0.24) | 16.12 (0.50) | <.0001 |
| Missing | 1,888 | 352 | 1,536 |  |
| Follow-up time, years | 19.98 (0.28) | 20.33 (0.26) | 18.92 (0.42) | <.0001 |
| Age, years | 54.66 (0.45) | 53.93 (0.48) | 56.87 (0.53) | <.0001 |
| Pack-years | 14.60 (0.43) | 14.26 (0.47) | 15.81 (0.84) | 0.0941 |
| Missing | 359 | 0 | 359 |  |
| Serum cotinine, ng/mL | 70.09 (2.50) | 70.24 (2.61) | 68.98 (5.91) | 0.8371 |
| Missing | 1,389 | 0 | 1,389 |  |
| Body mass index, kg/m^2^ | 27.32 (0.13) | 27.36 (0.14) | 27.13 (0.22) | 0.3287 |
| Missing | 822 | 0 | 822 |  |
| **Categorical variables, n (%)** | | | | |
| Sex |  |  |  | 0.0002 |
| Male | 3,975 (45.47%) | 2,827 (46.74%) | 1,148 (41.65%) |  |
| Female | 4,817 (54.53%) | 3,390 (53.26%) | 1.427 (58.35%) |  |
| Race/ethnicity |  |  |  | <.0001 |
| Non-Hispanic White | 4,394 (81.37%) | 3,181 (82.59%) | 1,213 (77.71%) |  |
| Non-Hispanic Black | 2,077 (8.93%) | 1,416 (8.15%) | 661 (11.28%) |  |
| Mexican American | 1,982 (3.52%) | 1,376 (3.39%) | 606 (3.90%) |  |
| Other | 339 (6.18%) | 244 (5.87%) | 95 (7.12%) |  |
| Education |  |  |  | <.0001 |
| Less than high school | 3,823 (26.82%) | 2,545 (24.69%) | 1,278 (33.46%) |  |
| High school graduate | 3,696 (51.19%) | 2,779 (52.34%) | 917 (47.62%) |  |
| College and above | 1,195 (21.98%) | 893 (22.97%) | 302 (18.92%) |  |
| Missing | 78 | 0 | 78 |  |
| Income-poverty ratio |  |  |  | 0.004 |
| <=1 | 1,452 (9.01%) | 1,104 (8.40%) | 348 (11.59%) |  |
| >1 | 6,458 (90.99%) | 5,113 (91.60%) | 1,345 (88.41%) |  |
| Missing | 882 | 0 | 882 |  |
| Smoking status |  |  |  | <.0001 |
| Never | 4,010 (42.44%) | 2,953 (44.05%) | 1,057 (37.59%) |  |
| Former | 2,793 (33.63%) | 1,949 (33.35%) | 844 (34.47%) |  |
| Current | 1.982 (23.93%) | 1,315 (22.60%) | 667 (27.95%) |  |
| Missing | 7 | 0 | 7 |  |
| Alcohol consumption |  |  |  | 0.5735 |
| 0 drink/day | 6,639 (86.98%) | 5,503 (87.12%) | 1,136 (86.02%) |  |
| ≥1 drink/day | 859 (13.02%) | 714 (12.88%) | 145 (13.98%) |  |
| Missing | 1,294 | 0 | 1,294 |  |

Note: Survey-weighted means and standard errors were calculated for continuous variables. Unweighted frequencies and survey-weighted percentages were calculated for categorical variables. Survey-weighted t-tests were used to test continuous variables, and survey-weighted Chi-square tests were used for categorical variables.

**Table S5.** Associations between lead and incident AD and all-cause dementia in NHANES-III.

|  | **Quartiles of lead concentrations^a^** | | | | | **Continuous lead concentrations** | |
| --- | --- | --- | --- | --- | --- | --- | --- |
|  | **Quartile 1** | **Quartile 2** | **Quartile 3** | **Quartile 4** | **P for trend** | **Per doubling** | **P** |
| **Association between lead and AD** | | | | | | | |
| Blood lead, N = 6,217 | | | | | | | |
| Range (µg/dL) | 0.7-1.9 | 2.0-3.1 | 3.2-4.7 | 4.8-52.9 |  |  |  |
| #Cases/Participants | 135/1,268 | 220/1,540 | 168/1,561 | 234/1,848 |  |  |  |
| HR (95% CI) | Ref | 1.34 (1.05, 1.70) | 0.98 (0.76, 1.27) | 1.15 (0.89, 1.49) | 0.94 | 1.01 (0.92, 1.11) | 0.86 |
| Estimated patella lead, N = 5,865 | | | | | | | |
| Range (µg/g) | 8.38-15.24 | 15.25-19.64 | 19.64-26.36 | 26.36-81.38 |  |  |  |
| #Cases/Participants | 17/938 | 64/1,191 | 195/1,568 | 429/2,168 |  |  |  |
| HR (95% CI) | Ref | 1.39 (0.81, 2.38) | 1.35 (0.76, 2.40) | 1.28 (0.67, 2.42) | 0.99 | 0.77 (0.53, 1.12) | 0.33 |
| Estimated tibia lead, N = 5,865 | | | | | | | |
| Range (µg/g) | 0-7.24 | 7.24-11.63 | 11.63-17.88 | 17.88-57.07 |  |  |  |
| #Cases/Participants | 14/900 | 48/1,189 | 181/1,577 | 462/2,199 |  |  |  |
| HR (95% CI) | Ref | 1.01 (0.56, 1.83) | 0.98 (0.51, 1.90) | 0.88 (0.41, 1.87) | 0.59 | 1.02 (0.84, 1.25) | 0.75 |
| **Association between lead and all-cause dementia** | | | | | | | |
| Blood lead, N = 5,865 | | | | | | | |
| Range (µg/dL) | 0.7-1.9 | 2.0-3.1 | 3.2-4.7 | 4.8-52.9 |  |  |  |
| #Cases/Participants | 265/1,268 | 391/1,540 | 386/1,561 | 501/1,848 |  |  |  |
| HR (95% CI) | Ref | 1.12 (0.94, 1.34) | 1.18 (0.98, 1.41) | 1.11 (0.92, 1.33) | 0.32 | 1.03 (0.96, 1.10) | 0.41 |
| Estimated patella lead, N = 5,865 | | | | | | | |
| Range (µg/g) | 8.38-15.24 | 15.25-19.64 | 19.64-26.36 | 26.36-81.38 |  |  |  |
| #Cases/Participants | 37/938 | 127/1,191 | 393/1,568 | 881/2,168 |  |  |  |
| HR (95% CI) | Ref | 1.49 (1.04, 2.13) | 1.73 (1.18, 2.54) | 1.67 (1.09, 2.56) | 0.24 | 0.96 (0.74, 1.23) | 0.77 |
| Estimated tibia lead, N = 5,865 | | | | | | | |
| Range (µg/g) | 0-7.24 | 7.24-11.63 | 11.63-17.88 | 17.88-57.07 |  |  |  |
| #Cases/Participants | 27/900 | 104/1,189 | 377/1,577 | 930/2,199 |  |  |  |
| HR (95% CI) | Ref | 1.74 (1.14, 2.66) | 2.02 (1.26, 3.22) | 1.87 (1.10, 3.19) | 0.44 | 1.19 (0.96, 1.46) | 0.11 |

AD: Alzheimer disease. All models were adjusted for age, age^2^, sex, race/ethnicity, education, income-poverty ratio, smoking status, pack-year, serum cotinine, alcohol consumption, and body mass index.

^a^ Quartiles of lead concentrations were based on survey-weighted distributions.

**Table S6.** Population attributable fractions for AD and all-cause dementia in NHANES-III.

| **PAF (95% CI)** | **Blood lead** | **Estimated patella lead** | **Estimated tibia lead** |
| --- | --- | --- | --- |
| Reference concentrations (25^th^ percentile) | 1.9 µg/dL | 15.2 µg/g | 7.2 µg/g |
| **AD** | -1% (-12%, 10%) | -12% (-42%, 16%) | 1% (-11%, 13%) |
| **All-cause dementia** | 0.4% (-6%, 7%) | -3% (-21%, 13%) | 11% (-6%, 26%) |

PAF: population attributable fraction; AD: Alzheimer disease. PAFs were derived from survey-weighted Cox proportional hazards models utilizing log_2_-transformed lead concentrations. Calculations compared observed exposure distributions to the 25^th^ percentile of each lead concentration as the theoretical low risk exposure distributions.

**Table S7.** Effect modification of associations of lead exposure with AD and all-cause dementia by sex in continuous NHANES 1999-2016.

|  | **Quartiles of lead concentrations^a^** | | | | | **Continuous lead concentrations** | |
| --- | --- | --- | --- | --- | --- | --- | --- |
|  | **Quartile 1** | **Quartile 2** | **Quartile 3** | **Quartile 4** | **P for interaction** | **Per doubling** | **P for interaction** |
| **Association between lead and AD** | | | | | | | |
| Blood lead | | | | | | | |
| Female | Ref | 0.92 (0.64, 1.32) | 0.79 (0.54, 1.14) | 0.69 (0.44, 1.08) | 0.17 | 0.83 (0.68, 1.00) | 0.08 |
| Male | Ref | 1.97 (0.95, 4.11) | 1.22 (0.63, 2.39) | 1.62 (0.79, 3.31) |  | 1.07 (0.89, 1.29) |  |
| Estimated patella lead^b^ | | | | | | | |
| Female | Ref | 2.57 (1.02, 6.49) | 2.43 (0.96, 6.16) | 2.64 (0.97, 7.17) | 0.22 | 1.49 (0.82, 2.71) | 0.34 |
| Male | Ref | 1.96 (0.64, 6.04) | 3.57 (1.34, 9.51) | 3.67 (1.16, 11.61) |  | 2.03 (1.12, 3.66) |  |
| Estimated tibia lead^b^ | | | | | | | |
| Female | Ref | 1.44 (0.68, 3.07) | 1.20 (0.52, 2.78) | 1.00 (0.40, 2.48) | 0.08 | 0.87 (0.47, 1.64) | 0.13 |
| Male | Ref | 0.91 (0.26, 3.24) | 1.42 (0.36, 5.59) | 1.48 (0.34, 6.56) |  | 1.43 (0.72, 2.82) |  |
| **Association between lead and all-cause dementia** | | | | | | | |
| Blood lead | | | | | | | |
| Female | Ref | 0.84 (0.65, 1.10) | 0.93 (0.71, 1.22) | 0.92 (0.69, 1.22) | 0.82 | 0.96 (0.84, 1.10) | 0.32 |
| Male | Ref | 1.03 (0.60, 1.75) | 0.77 (0.47, 1.26) | 1.01 (0.62, 1.65) |  | 1.05 (0.91, 1.22) |  |
| Estimated patella lead^b^ | | | | | | | |
| Female | Ref | 2.09 (1.11, 3.95) | 2.06 (1.08, 3.94) | 2.73 (1.40, 5.30) | 0.90 | 1.97 (1.28, 3.04) | 0.47 |
| Male | Ref | 0.88 (0.41, 1.92) | 1.33 (0.74, 2.40) | 1.66 (0.84, 3.28) |  | 1.69 (1.04, 2.74) |  |
| Estimated tibia lead^b^ | | | | | | | |
| Female | Ref | 1.29 (0.74, 2.26) | 1.27 (0.68, 2.38) | 1.46 (0.79, 2.71) | 0.98 | 1.69 (1.13, 2.51) | 0.67 |
| Male | Ref | 1.08 (0.56, 2.11) | 1.29 (0.65, 2.57) | 1.34 (0.59, 3.05) |  | 1.55 (0.95, 2.55) |  |

AD: Alzheimer disease. All models were adjusted for age, age^2^, sex, race/ethnicity, education, income-poverty ratio, smoking status, pack-year, serum cotinine, alcohol consumption, and body mass index. Multiplicative interaction terms between lead and sex were also included in the models.

^a^ Quartiles of lead concentrations were based on survey-weighted distributions.

^b^ Bone lead was available in NHANES 1999-2014.

**Table S8.** Effect modification of associations of lead exposure with AD and all-cause dementia by sex in NHANES-III

|  | **Quartiles of lead concentrations^a^** | | | | | **Continuous lead concentrations** | |
| --- | --- | --- | --- | --- | --- | --- | --- |
|  | **Quartile 1** | **Quartile 2** | **Quartile 3** | **Quartile 4** | **P for interaction** | **Per doubling** | **P for interaction** |
| **Association between lead and AD** | | | | | | | |
| Blood lead | | | | | | | |
| Female | Ref | 1.32 (0.82, 2.12) | 1.04 (0.63, 1.72) | 1.10 (0.74, 1.62) | 0.93 | 0.99 (0.86, 1.13) | 0.57 |
| Male | Ref | 1.37 (0.80, 2.35) | 0.88 (0.52, 1.51) | 1.22 (0.71, 2.09) |  | 1.06 (0.86, 1.32) |  |
| Estimated patella lead | | | | | | | |
| Female | Ref | 1.11 (0.41, 3.03) | 1.02 (0.35, 3.00) | 0.94 (0.32, 2.73) | 0.19 | 0.69 (0.41, 1.17) | 0.22 |
| Male | Ref | 3.54 (1.15, 10.93) | 3.80 (0.89, 16.27) | 3.72 (0.88, 15.75) |  | 0.89 (0.49, 1.60) |  |
| Estimated tibia lead | | | | | | | |
| Female | Ref | 0.91 (0.29, 2.84) | 0.84 (0.23, 3.01) | 0.73 (0.19, 2.90) | 0.29 | 1.02 (0.89, 1.15) | 0.40 |
| Male | Ref | 1.60 (0.51, 5.10) | 1.76 (0.55, 5.57) | 1.66 (0.47, 5.83) |  | 1.16 (0.87, 1.54) |  |
| **Association between lead and all-cause dementia** | | | | | | | |
| Blood lead | | | | | | | |
| Female | Ref | 1.12 (0.81, 1.55) | 1.31 (0.99, 1.73) | 1.15 (0.86, 1.53) | 0.22 | 1.05 (0.96, 1.15) | 0.48 |
| Male | Ref | 1.07 (0.68, 1.67) | 0.93 (0.63, 1.37) | 0.96 (0.66, 1.39) |  | 0.99 (0.87, 1.11) |  |
| Estimated patella lead | | | | | | | |
| Female | Ref | 1.33 (0.67, 2.63) | 1.45 (0.73, 2.91) | 1.39 (0.67, 2.88) | 0.35 | 0.92 (0.67. 1.26) | 0.62 |
| Male | Ref | 2.04 (0.82, 5.08) | 2.61 (0.99, 6.92) | 2.52 (0.91, 7.03) |  | 1.00 (0.68, 1.47) |  |
| Estimated tibia lead | | | | | | | |
| Female | Ref | 1.63 (0.73, 3.62) | 1.70 (0.72, 4.01) | 1.50 (0.59, 3.84) | 0.07 | 1.16 (0.96, 1.41) | 0.46 |
| Male | Ref | 2.64 (0.87, 7.98) | 3.83 (1.29, 11.40) | 3.87 (1.21, 12.32) |  | 1.29 (0.94, 1.76) |  |

AD: Alzheimer disease. All models were adjusted for age, age^2^, sex, race/ethnicity, education, income-poverty ratio, smoking status, pack-year, serum cotinine, alcohol consumption, and body mass index. Multiplicative interaction terms between lead and sex were also included in the models.

^a^ Quartiles of lead concentrations were based on survey-weighted distributions.

**Table S9**. Associations between bone lead and AD and all-cause dementia after further adjusting for blood lead in continuous NHANES 1999-2014.

|  | **Quartiles of lead concentrations^a^** | | | | | **Continuous lead concentrations** | |
| --- | --- | --- | --- | --- | --- | --- | --- |
|  | **Quartile 1** | **Quartile 2** | **Quartile 3** | **Quartile 4** | **P for trend** | **Per doubling** | **P** |
| **Association between bone lead and AD** | | | | | | | |
| Estimated patella lead | Ref | 2.47 (1.33, 4.60) | 2.86 (1.44, 5.70) | 3.14 (1.43, 6.89) | 0.12 | 2.41 (1.23, 4.70) | 0.02 |
| Estimated tibia lead | Ref | 1.26 (0.70, 2.29) | 1.22 (0.62, 2.39) | 1.10 (0.49, 2.45) | 0.79 | 1.31 (0.57, 2.99) | 0.61 |
| **Association between bone lead and all-cause dementia** | | | | | | | |
| Estimated patella lead | Ref | 1.47 (1.01, 2.14) | 1.64 (1.07, 2.50) | 2.09 (1.28, 3.40) | 0.02 | 1.94 (1.23, 3.06) | 0.03 |
| Estimated tibia lead | Ref | 1.17 (0.79, 1.74) | 1.21 (0.77, 1.90) | 1.28 (0.75, 2.21) | 0.50 | 2.06 (1.17, 3.63) | 0.02 |

AD: Alzheimer disease. All models were adjusted for blood lead, age, age^2^, sex, race/ethnicity, education, income-poverty ratio, smoking status, pack-year, serum cotinine, alcohol consumption, and body mass index.

^a^ Quartiles of lead concentrations were based on survey-weighted distributions.

**Table S10**. Associations between bone lead and AD and all-cause dementia after further adjusting for blood lead in NHANES-III.

|  | **Quartiles of lead concentrations^a^** | | | | | **Continuous lead concentrations** | |
| --- | --- | --- | --- | --- | --- | --- | --- |
|  | **Quartile 1** | **Quartile 2** | **Quartile 3** | **Quartile 4** | **P for trend** | **Per doubling** | **P** |
| **Association between bone lead and AD** | | | | | | | |
| Estimated patella lead | Ref | 1.39 (0.80, 2.39) | 1.34 (0.74, 2.43) | 1.26 (0.64, 2.50) | 0.99 | 0.58 (0.34, 1.00) | 0.15 |
| Estimated tibia lead | Ref | 0.98 (0.53, 1.80) | 0.93 (0.46, 1.88) | 0.82 (0.36, 1.88) | 0.51 | 1.03 (0.81, 1.30) | 0.63 |
| **Association between bone lead and all-cause dementia** | | | | | | | |
| Estimated patella lead | Ref | 1.47 (1.02, 2.12) | 1.70 (1.14, 2.52) | 1.61 (1.02, 2.56) | 0.33 | 0.76 (0.53, 1.10) | 0.26 |
| Estimated tibia lead | Ref | 1.70 (1.11, 2.62) | 1.94 (1.18, 3.18) | 1.77 (0.98, 3.17) | 0.61 | 1.23 (0.90, 1.69) | 0.30 |

AD: Alzheimer disease. All models were adjusted for blood lead, age, age^2^, sex, race/ethnicity, education, income-poverty ratio, smoking status, pack-year, serum cotinine, alcohol consumption, and body mass index.

^a^ Quartiles of lead concentrations were based on survey-weighted distributions.

**Table S11**. Associations between lead and incident AD and all-cause dementia in continuous NHANES 1999-2016 using Fine-Gray competing risk regression models.

|  | **AD models** | | **All-cause dementia models** | |
| --- | --- | --- | --- | --- |
|  | **AD** | **Non-AD Mortality** | **All-cause dementia** | **Non-dementia mortality** |
| **Blood lead** | | | | |
| # events/total | 587/8,038 | 2,135/8,038 | 1,260/8,038 | 1,683/8,038 |
| Subdistribution hazard model | 0.99 (0.90, 1.10) | 1.04 (0.98, 1.10) | 1.09 (1.01, 1.17) | 1.03 (0.97, 1.10) |
| Cause-specific hazard model | 0.90 (0.80, 1.02) | 1.01 (0.94, 1.07) | 0.99 (0.92, 1.08) | 1.03 (0.96, 1.11) |
| **Estimated patella lead** | | | | |
| # events/total | 443/4,824 | 1,534/4,824 | 922/4,824 | 1,186/4,824 |
| Subdistribution hazard model | 1.26 (0.77, 2.06) | 1.34 (1.02, 1.75) | 1.48 (1.05, 2.08) | 1.31 (0.97, 1.77) |
| Cause-specific hazard model | 1.74 (1.00, 3.01) | 1.57 (1.19, 2.08) | 1.82 (1.26, 2.63) | 1.55 (1.13, 2.13) |
| **Estimated tibia lead** | | | | |
| # events/total | 443/4,824 | 1,534/4,824 | 922/4,824 | 1,186/4,824 |
| Subdistribution hazard model | 1.03 (0.65, 1.62) | 1.30 (1.00, 1.69) | 1.57 (1.14, 2.16) | 1.21 (0.90, 1.63) |
| Cause-specific hazard model | 1.03 (0.60, 1.78) | 1.28 (0.97, 1.67) | 1.63 (1.14, 2.34) | 1.19 (0.88, 1.61) |

AD: Alzheimer disease. All models were adjusted for age, age^2^, sex, race/ethnicity, education, income-poverty ratio, smoking status, pack-year, serum cotinine, alcohol consumption, and body mass index.

Subdistribution hazard model = Fine-Gray competing risk analysis. The competing risk outcome was coded as incident AD or all-cause dementia, mortality (competing risk), or censored.

Cause-specific hazard model = Cox proportional hazard model evaluating either AD/all-cause dementia or mortality as separate outcomes.

**Table S12**. Associations between lead and incident AD and all-cause dementia in NHANES-III using Fine-Gray competing risk regression models.

|  | **AD models** | | **All-cause dementia models** | |
| --- | --- | --- | --- | --- |
|  | **AD** | **Non-AD Mortality** | **All-cause dementia** | **Non-dementia mortality** |
| **Blood lead** | | | | |
| # events/total | 757/6,217 | 2,711/6,217 | 1,543/6,217 | 2,052/6,217 |
| Subdistribution hazard model | 0.94 (0.87, 1.02) | 1.08 (1.03, 1.13) | 0.99 (0.93, 1.05) | 1.08 (1.02, 1.14) |
| Cause-specific hazard model | 1.01 (0.92, 1.11) | 1.12 (1.07, 1.18) | 1.03 (0.96, 1.10) | 1.12 (1.06, 1.19) |
| **Estimated patella lead** | | | | |
| # events/total | 705/5,865 | 2,559/5,865 | 1,438/5,865 | 1,945/5,865 |
| Subdistribution hazard model | 0.72 (0.52, 1.00) | 1.30 (1.11, 1.54) | 0.97 (0.78, 1.21) | 1.22 (1.02, 1.47) |
| Cause-specific hazard model | 0.77 (0.53, 1.12) | 1.25 (1.05, 1.48) | 0.96 (0.74, 1.23) | 1.25 (1.03, 1.52) |
| **Estimated tibia lead** | | | | |
| # events/total | 705/5,865 | 2,559/5,865 | 1,438/5,865 | 1,945/5,865 |
| Subdistribution hazard model | 0.98 (0.92, 1.05) | 142 (1.22, 1.64) | 1.08 (0.96, 1.22) | 1.40 (1.19, 1.65) |
| Cause-specific hazard model | 1.02 (0.84, 1.25) | 1.38 (1.19, 1.60) | 1.19 (0.96, 1.46) | 1.33 (1.13, 1.57) |

AD: Alzheimer disease. All models were adjusted for age, age^2^, sex, race/ethnicity, education, income-poverty ratio, smoking status, pack-year, serum cotinine, alcohol consumption, and body mass index.

Subdistribution hazard model = Fine-Gray competing risk analysis. The competing risk outcome was coded as incident AD or all-cause dementia, mortality (competing risk), or censored.

Cause-specific hazard model = Cox proportional hazard model evaluating either AD/all-cause dementia or mortality as separate outcomes.

**Supplementary Figures**

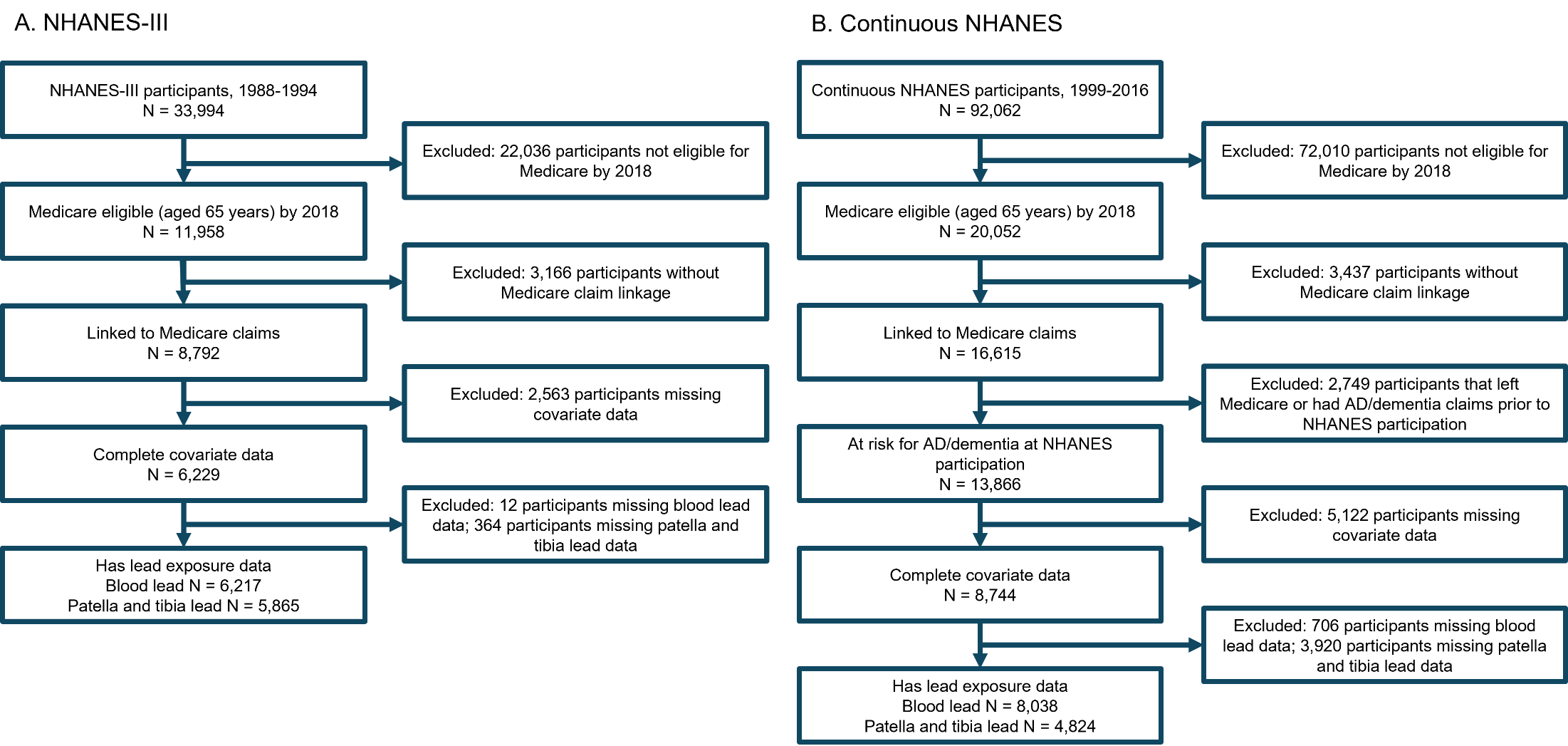

**Figure S1.** Flowcharts depicting participant inclusion/exclusion criteria for the two independent analytic samples, (A) NHANES-III and (B) continuous NHANES. AD: Alzheimer disease.

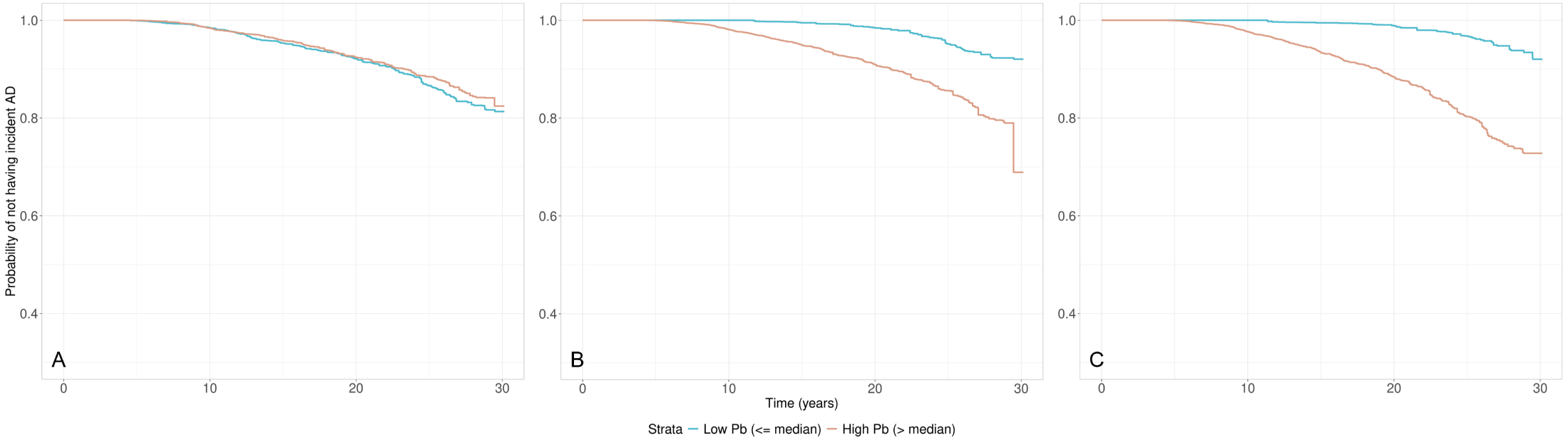

**Figure S2**. Kaplan-Meier (K-M) curves showing the probability of remaining free from Alzheimer disease (AD), stratified by median concentrations of (A) blood lead (3.1 µg/dL), (B) estimated patella lead (19.6 µg/g), and (C) estimated tibia lead (11.6 µg/g), in NHANES-III. The curves account for survey weights and inverse probability weighting based on key covariates including age, sex, race/ethnicity, education, income-poverty ratio, smoking status, pack-year, serum cotinine, alcohol consumption, and body mass index.

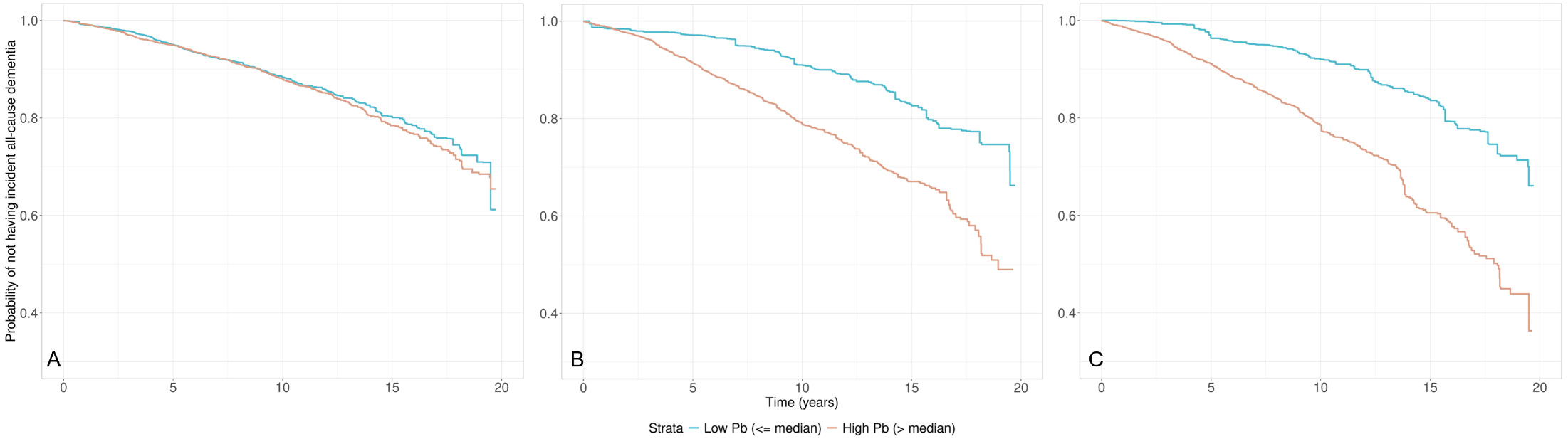

**Figure S3**. Kaplan-Meier (K-M) curves showing the probability of remaining free from all-cause dementia, stratified by median concentrations of (A) blood lead (1.7 µg/dL), (B) estimated patella lead (23.9 µg/g), and (C) estimated tibia lead (16.4 µg/g), in continuous NHANES. The curves account for survey weights and inverse probability weighting based on key covariates including age, sex, race/ethnicity, education, income-poverty ratio, smoking status, pack-year, serum cotinine, alcohol consumption, and body mass index.

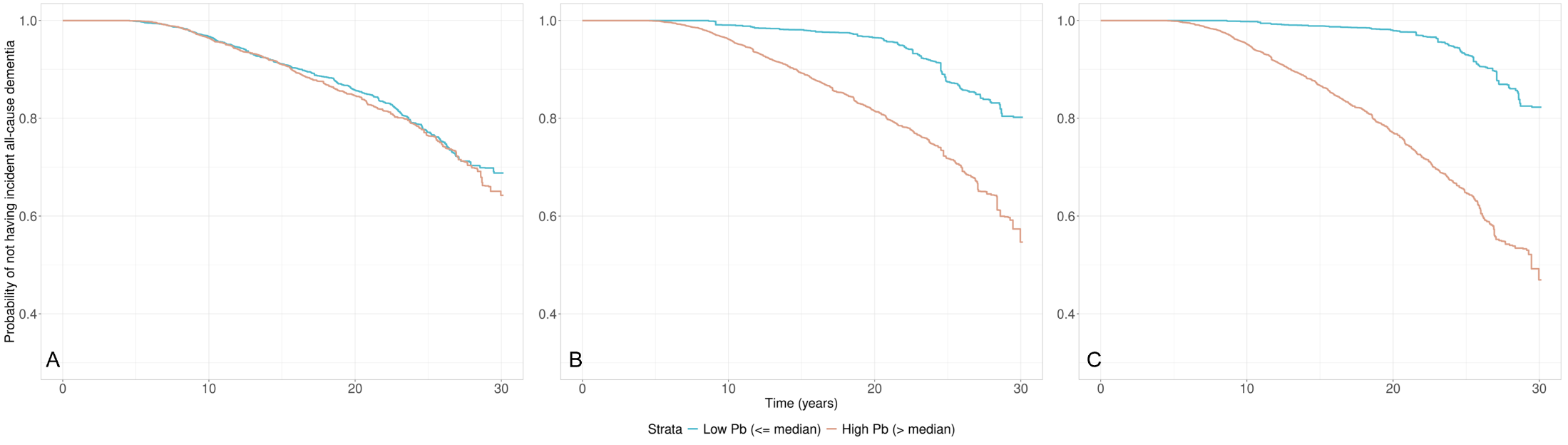

**Figure S4**. Kaplan-Meier (K-M) curves showing the probability of remaining free from all-cause dementia, stratified by median concentrations of (A) blood lead (3.1 µg/dL), (B) estimated patella lead (19.6 µg/g), and (C) estimated tibia lead (11.6 µg/g), in NHANES-III. The curves account for survey weights and inverse probability weighting based on key covariates including age, sex, race/ethnicity, education, income-poverty ratio, smoking status, pack-year, serum cotinine, alcohol consumption, and body mass index.
